# Employment status and its associated factor for patients 12 months after intensive care: Secondary analysis of the SMAP-HoPe-study

**DOI:** 10.1101/2021.12.01.21266910

**Authors:** Takeshi Unoki, Mio Kitayama, Hideaki Sakuramoto, Akira Ouchi, Tomoki Kuribara, Takako Yamaguchi, Sakura Uemura, Yuko Fukuda, Junpei Haruna, Takahiro Tsujimoto, Mayumi Hino, Yuko Shiba, Takumi Nagao, Masako Shirasaka, Yosuke Satoi, Miki Toyoshima, Yoshiki Masuda, on behalf of the SMAP-HoPe Study Project

## Abstract

Returning to work is a serious issue that affects patients who are being discharged from the intensive care unit (ICU). This study aimed to clarify the employment status and the perceived household financial status of ICU patients 12 months following discharge from the ICU. Additionally, a hypothesis of whether depressive symptoms were associated with subsequent unemployment status was tested. This study was a subgroup analysis using data from the published Survey of Multicenter Assessment with Postal questionnaire for Post-Intensive Care Syndrome (PICS) for Home Living Patients (the SMAP-HoPe study) in Japan. The patients included those who had a history of staying in the ICU for at least three nights and had been living at home for one year following discharge, between October 2019 and July 2020. We assessed employment status, subjective cognitive functions, household financial status, Hospital Anxiety and Depression Scale scores, and EuroQOL-5 dimensions of physical function at 12 months following intensive care. This study included 328 patients who were known to be employed prior to ICU admission. The median age was 64 (Interquartile Range [IQR] 52-72), and males were predominant (86%). Seventy-nine (24%) of those evaluated were unemployed. The number of patients who reported worsened financial status was significantly higher in the unemployed group. (p<.01) Multivariate analysis showed that higher age (Odds Ratio [OR]: 1.06, 95% Confidence Interval [CI]: 1.03-1.08]) and severity of depressive symptoms (OR: 1.13 [95% CI: 1.05-1.23]) were independent factors for employment status after 12 months from being discharged from the ICU. These factors were determined to be significant even after adjusting for sex, physical function, and cognitive function. We found that one-fourth of our patients who had been employed prior to ICU admission were subsequently unemployed 12 months following ICU discharge. Additionally, depressive symptoms were associated with unemployment status. The government and the local municipalities should provide medical and financial support to such patients. Additionally, community support for such patients is warranted.

## Introduction

The problem of patients returning to work following discharge from the intensive care unit (ICU) is a serious issue. A systematic review and meta-analysis, including 52 studies, of returning to work among previously critically ill patients indicated that delayed return to work and unemployment were common and persistent problems[1]. These studies showed that 36% of patients were subsequently unemployed at 12 months following ICU admission. This change in employment status shall have a corresponding effect on the household income. A study suggested that 30% of patients discharged from the ICU had a decline in household income even after 6 months to one year following intensive care admission in the United Kingdom[2].

National employment and disability policies could contribute to resumption of work productivity among those discharged from the ICU. Su et al. [3] revealed that the disability policies of each country were related to resumption of work productivity. Thus, it is worth evaluating how those discharged from the ICU fare in terms of employment in each country. Notably, there was a prior study examining the employment status of those discharged from the ICU in Japan.[4] They showed that among 33 patients discharged from the ICU, 26 (83.9%) were able to return to work six months following ICU admission. However, the precision was insufficient due to a small sample size. For enhanced understanding of employment status among those discharged from the ICU, a study with a larger sample size was therefore warranted.

The factors associated with the unemployment of those discharged from the ICU had been examined in previous studies. Higher age and female [5], cognitive function [6], depressive symptoms [7], physical disability and educational level [8] had been suggested to influence unemployment status in those discharged from the ICU. However, these factors are not consistent between studies[1]. Among the general population, absence due to all sickness and depression are key factors influencing a longer duration until return to work [9].

The primary objective of our study was to elucidate the employment status and perceived household financial status among those discharged from the ICU 12 months following discharge. The secondary objective was to test a hypothesis of whether depressive symptoms were associated with subsequent employment status following discharge from intensive care.

## Materials and methods

### Study design

The present study was a sub analysis of the the Survey of Multicenter Assessment with Postal questionnaire for Post-Intensive Care Syndrome (PICS) for Home Living Patients (SMAP-HoPe) study [10] and focused on employment status following ICU discharge and the factors that affect it. This analysis was nested in the SMAP-HoPe study [10]. In summary of SMAP-HoPe study, we conducted an ambidirectional study for patients living at home 12 months following ICU discharge. We sent postal surveys on post-intensive care syndrome and employment status. Data from when the patients were admitted were obtained for retrospective analysis. Detailed methods were provided elsewhere in a distinct publication [10].

### Setting

Twelve ICUs in Japan were included in this study. The detailed characteristics of each ICU included in the study were previously reported in the original study [10].

### Participants

The patients included those who had stayed in the ICU for at least three nights and had been living at home for one year following ICU discharge between October 2019 and July 2020. The number of subjects in the previously published study of which this is a subgroup analysis was 754 patients; the response rate was 91.1%. In this secondary analysis focusing on employment status, only patients aged 18 years or over and who had been working prior to admission were included.

### Variables/Instruments/Data source

Patients’ characteristics including age, sex, diagnosis, APACHE II score, pre-existing disease, and length of ICU stay were retrospectively collected by medical chart review. Status after 12 months following discharge from intensive care was collected through postal questionnaires, which explored work status, cognitive function, and Euro-QOL-5D-5L (EQ-5D-5L). [11][11] EQ-5D-5L is a validated questionnaire [11] and Japanese version is available upon request to the EuroQOL group (https://euroqol.org). Work status prior to ICU admission and present working status were classified according to the following scheme: unemployed, self-employed, employed part-time, and employed full-time. Additionally, we inquired about whether household finances changed compared with the period prior to ICU admission. The respondents selected between a three-point scale of “worse,” “no change,” or “better.” Cognitive function was measured using two simple questions that we developed, due to lack of valid instruments for self-administration. One was “Do you think your memory function was impaired compared with before ICU admission?” Another was “Do you think your concentration function was impaired compared with before ICU admission?” Four-point Likert scales, from not at all (0), sometimes (1), frequently (2), and very frequently (3) were the acceptable responses. If the subject responds “very frequently” or “frequently” for either of the two questions, we define them to have cognitive dysfunction. Usual activities were measured by part of question from EQ-5D-5L. The response consisted of five levels, from having “no problems” performing usual activities to being “unable to do” their usual activities. We defined responses other than “no problems” or “slight problems” as evidence of physical dysfunction. Severity of depression was measured by the Hospital Anxiety and Depression Scale (HADS) [12]. HADS consists of depression and anxiety subscales, and each subscale has seven components, which were rated on a scale from 0 to 3. Subscales of depression are graded from 0 to 21. Previous studies demonstrated that the Japanese version of HADS had good reliability and validity [13]. We used Japanese version of HADS [13].

### Bias

This study was a postal survey and selection bias may exist; however, selection bias was minimum, given the high rate of response from the original study from which this one was nested. Additionally, a state of emergency was declared by the government due to the coronavirus disease 2019 (COVID-19) pandemic in April 2020, which overlapped with the study period. This declaration may have had an impact on employment. This bias was assessed through sensitivity analysis. Furthermore, in many Japanese companies, the compulsory age of retirement was generally between 60 and 65 years. The compulsory retirement, other than the episodes of intensive care admission, may influence our results. Thus, we accordingly conducted sensitivity analysis to evaluate its effect on our findings.

### Sensitivity Analysis

We conducted sensitivity analyses to confirm whether our findings were robust. First, we calculated the percentage of unemployment after excluding the subjects who responded within the duration of the COVID-19 pandemic; employment status after 12 months following ICU discharge may be influenced by the COVID-19 pandemic. Nevertheless employment status before ICU admission was collected between September 2018 and July 2019. We defined the period after April 2020 as the start of the COVID-19 pandemic because a state of emergency had been declared across the country on April 16, 2020. We calculated the proportion of the unemployed after excluding from the analysis the subjects who posted their responses during this period. Second, 60 years old was the beginning of eligibility for retirement from employment [14]. Thus, we performed multivariable analysis after excluding the subjects who were 60 years old and above and reviewed the impact of retirement age on our findings.

### Statistical analysis

First, descriptive statistical analyses were performed. Continuous variables were expressed as median [interquartile range (IQR)]. Wilcoxon rank-sum test was used for the comparison of continuous variables. Chi-square test was used for categorical variables. We calculated the proportion of unemployed subjects stratified by age. For multivariable analysis, a multilevel generalized linear model (GLM) with a binomial distribution and log link was performed. We pre-defined covariates consisting of work status before ICU admission, age [5], sex [5], cognitive dysfunction [6], physical dysfunction [7], and severity of depression [8] based on previous studies. Statistical analysis was used for STATA IC ver. 16 (Statacorp, TX) and R 4.0.2 (The R foundation for Statistical Computing), and the significance difference was set < .05.

### Ethical considerations

Human Research Ethics Committee approval was obtained. (Sapporo City University, Sapporo, Hokkaido, Japan, approval number: 1927-1) Additionally, ethical approval was obtained from all sites for the original studies. Detailed informed consent process was provided in the original study [10].

## Results

### Participants

Fig 1 displays a flow diagram of study enrollment. The SMAP-HoPe study had 754 subjects. We excluded 425 subjects who were unemployed before ICU admission. One patient who did not provide an answer to the question of cognitive function was excluded. Thus, 328 subjects were analyzed in this study.

**Fig 1.**
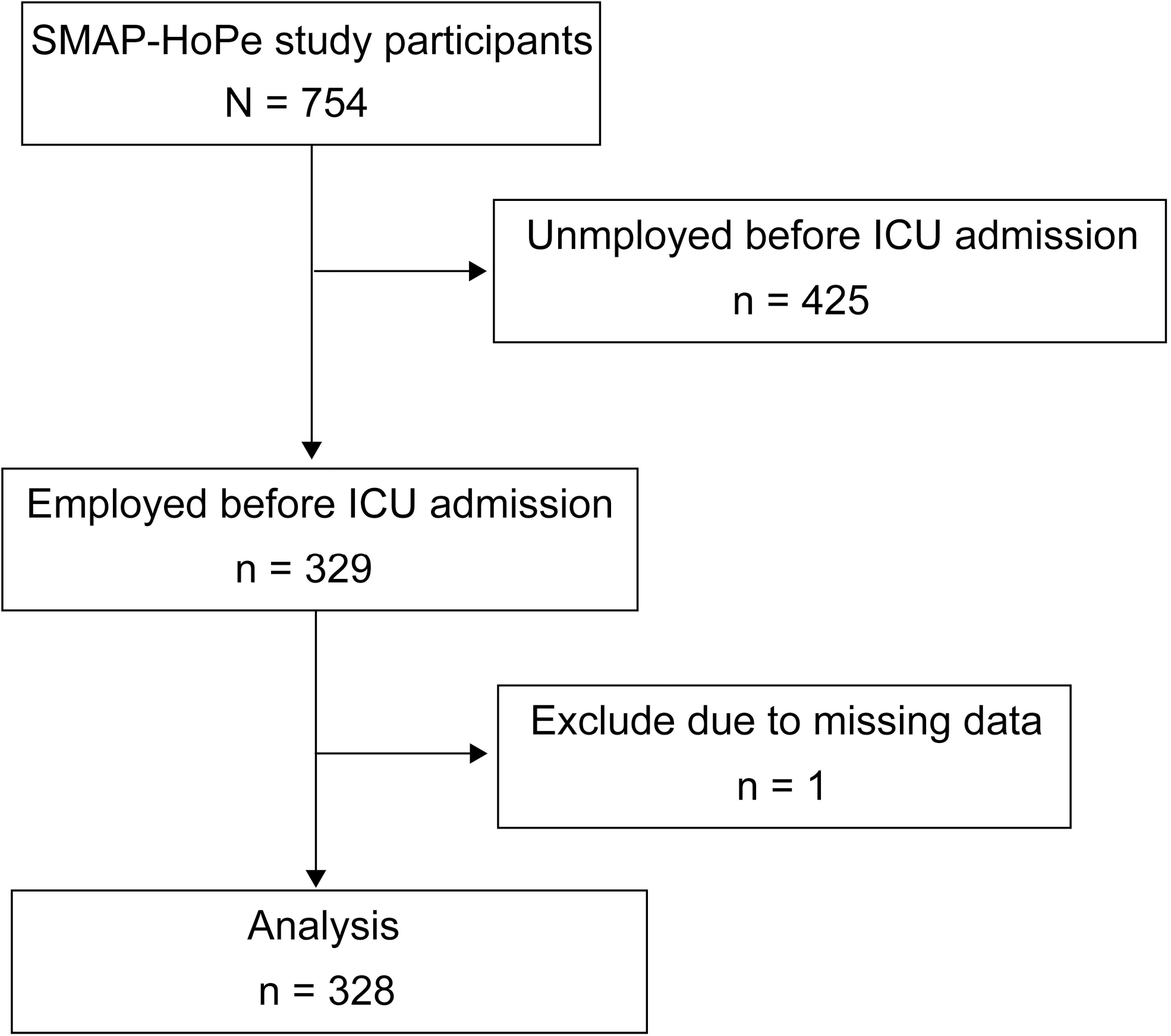
Patient recruitment scheme.

### Population of the study

The characteristics of the population are shown in Table 1. The median age of the population was 64 [IQR: 52-72] years. The number of subjects in each age group was shown in S1 Table. Male was dominant (86%). Approximately 41.2% of the subjects were admitted to the ICU after undergoing elective surgery. The number of subjects receiving mechanical ventilation was 219 (66.8%), and the median duration of mechanical ventilation were 2 days. Of 328 employed subjects before ICU admission, the most frequent employment status was part-time employed in 125 (50.2%) following full-time employed in 86 (34.5%)

**Table 1.** Demographic characteristics of employed and unemployed subjects from the SMAP-HoPe study.

| Variables | Participants n=328 | Employed n=249 | Unemployed n=79 | p-value |
| --- | --- | --- | --- | --- |
| Age (years), median [IQR] | 64 [52-72] | 62 [50-71] | 69 [62-75] | <.001 |
| Female, n (%) | 46 (14.0) | 32 (12.9) | 14 (17.7) | .271 |

| Types of admission, n (%) |  |  |  |  |
| --- | --- | --- | --- | --- |
| Elective surgery | 149 (41.2) | 109 (43.8) | 37 (46.2) | .697 |
| Unscheduled admission | 182 (55.5) | 140 (56.2) | 42 (53.2) | .697 |
| Unscheduled surgery | 49 (14.9) | 35 (14.1) | 14 (17.7) | .469 |
| Reason for ICU admission, n (%) |  |  |  |  |
| CV surgery | 122 (37.2) | 87 (34.9) | 35 (44.3) | .183 |
| CHF/AMI/Arrhy | 57 (17.4) | 46 (18.5) | 11 (13.9) |  |
| Sepsis | 34 (10.4) | 21 (8.4) | 13 (16.5) |  |
| Abdominal surgery | 30 (9.1) | 25 (10.0) | 5 (6.3) |  |
| Other surgery | 14 (4.3) | 9 (3.6) | 5 (6.3) |  |
| Trauma | 14 (4.3) | 13 (5.2) | 1 (1.3) |  |
| ENT surgery | 12 (3.7) | 10 (4.0) | 2 (2.5) |  |
| Aortic dissection (non-operative) | 12 (3.7) | 11 (4.4) | 1 (1.3) |  |
| Acute renal failure | 12 (3.7) | 9 (3.6) | 3 (3.8) |  |
| Others | 21 (6.4) | 18 (7.2) | 3 (3.8) |  |
| Employment status before ICU admission, n (%) |  |  |  |  |
| Full time | 101 (30.8) | 86 (34.5) | 15 (19.0) | <.001 |
| **Part time** | 156 (47.6) | 125 (50.2) | 31 (39.2) |  |
| **Self** | 71 (21.6) | 38 (15.3) | 33 (41.8) |  |
| **APACHE II , median [IQR]** | 14 [10-19] | 13 [9-19] | 16 [13-19] | .001 |
| **MV use, n (%)** | 219 (66.8) | 157 (63.1) | 62 (78.5) | .013 |
| **MV (days), median [IQR]** | 2.0 [0.0-3.0] | 2.0 [0.0-3.0] | 2.0 [1.0-3.0] | .041 |
| **Psychological history, n (%)** | 4 (1.2) | 4 (1.6) | 0 (0.0) | .576 |
| **Delirium (days), median [IQR]** | 0.0 [0.0-1.0] | 0.0 [0.0-1.0] | 0.0 [0.0-2.0] | .094 |
| **ICU LOS (days), median [IQR]** | 5.0 [4.0-7.0] | 5.0 [4.0-7.0] | 5.0 [4.0-7.0] | .293 |
| **Hospital LOS (days), median [IQR]** | 26.0 [18.0-36.0] | 25.0 [18.0-34.0] | 27.0 [19.0-46.5] | .192 |

IQR, interquartile range; CV, cardiovascular; CHF/AMI/arrhy, congestive heart failure/ acute myocardial infarction/ arrhythmia; ENT, ear nose throat; APACHE II , Acute Physiology and Chronic Health Evaluation II ; MV, mechanical ventilation; ICU, intensive care unit; LOS, length of stay; SMAP-HoPe: Survey of Multicenter Assessment with Postal questionnaire for Post-Intensive Care Syndrome (PICS) for Home Living Patients

### Characteristics between employed and unemployed subjects

Among 328 subjects who were previously employed, 79 (24.1%, 95% CI 19.7-29.1) were unemployed at 12 months following ICU discharge. A comparison of patient characteristics between the employed and the unemployed subjects is shown in Table 1. The median age of those who were employed was significantly lower than that of those who were unemployed (62 [50-71] vs 69 [62-75], p<0.01, respectively). The median APACHE II score among employed subjects was lower than that among unemployed subjects (13 [9-19] vs 16 [13-19], p<0.001, respectively).

### Employment status

Fig 2 shows the employment status between the period prior to ICU admission and 12 months following ICU discharge. Prior to ICU admission, the full-time employees comprised 101 subjects. Of those, 15 (14.9%) of them were unemployed at 12 months following discharge from the ICU. Additionally, of 156 subjects with part-time employment, 31 subjects were unemployed at 12 months following ICU discharge.

**Figure 2.**
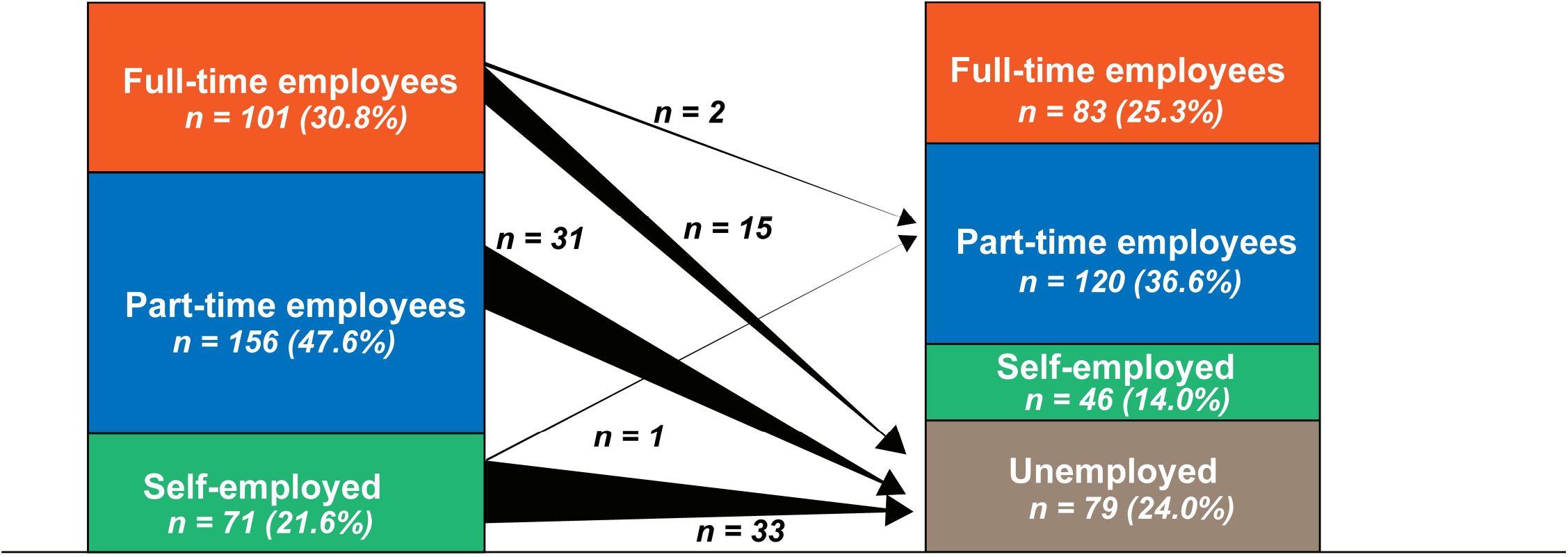
Change in the work status between before ICU admission and 12 months after ICU discharge.

### Household financial status

Fig 3 showed the proportion of respondents who perceived lower household financial status ; this figure distinguishes between employed and unemployed subjects at 12 months following ICU discharge. Among the subjects with an unemployed status, over half of them perceived their financial status to be worse compared with that in the period prior to ICU admission (p<.001).

**Figure 3.**
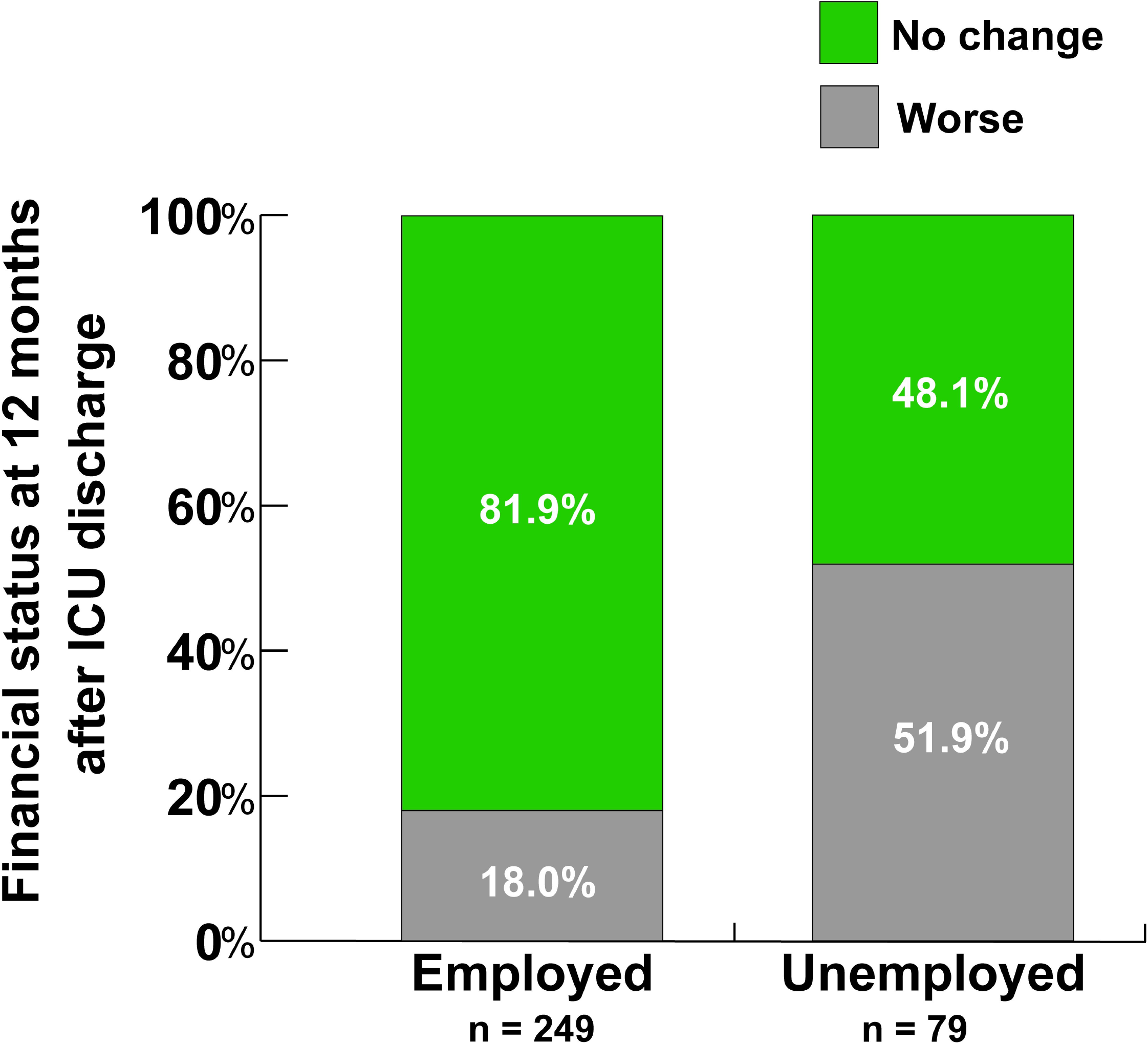
Perceived change in household financial status before and after ICU admission. The number of patients who reported that their household financial status had worsened was significantly higher among the unemployed 12 months after ICU discharge. (p< .001)

### Multivariable analysis

In multilevel GLM, it showed that higher age, part-time or self-employed status prior to ICU admission, and higher depressive symptoms were independent factors for being unemployed at 12 months following admission. The results of the multivariable analysis are shown in Table 2.

**Table 2.**
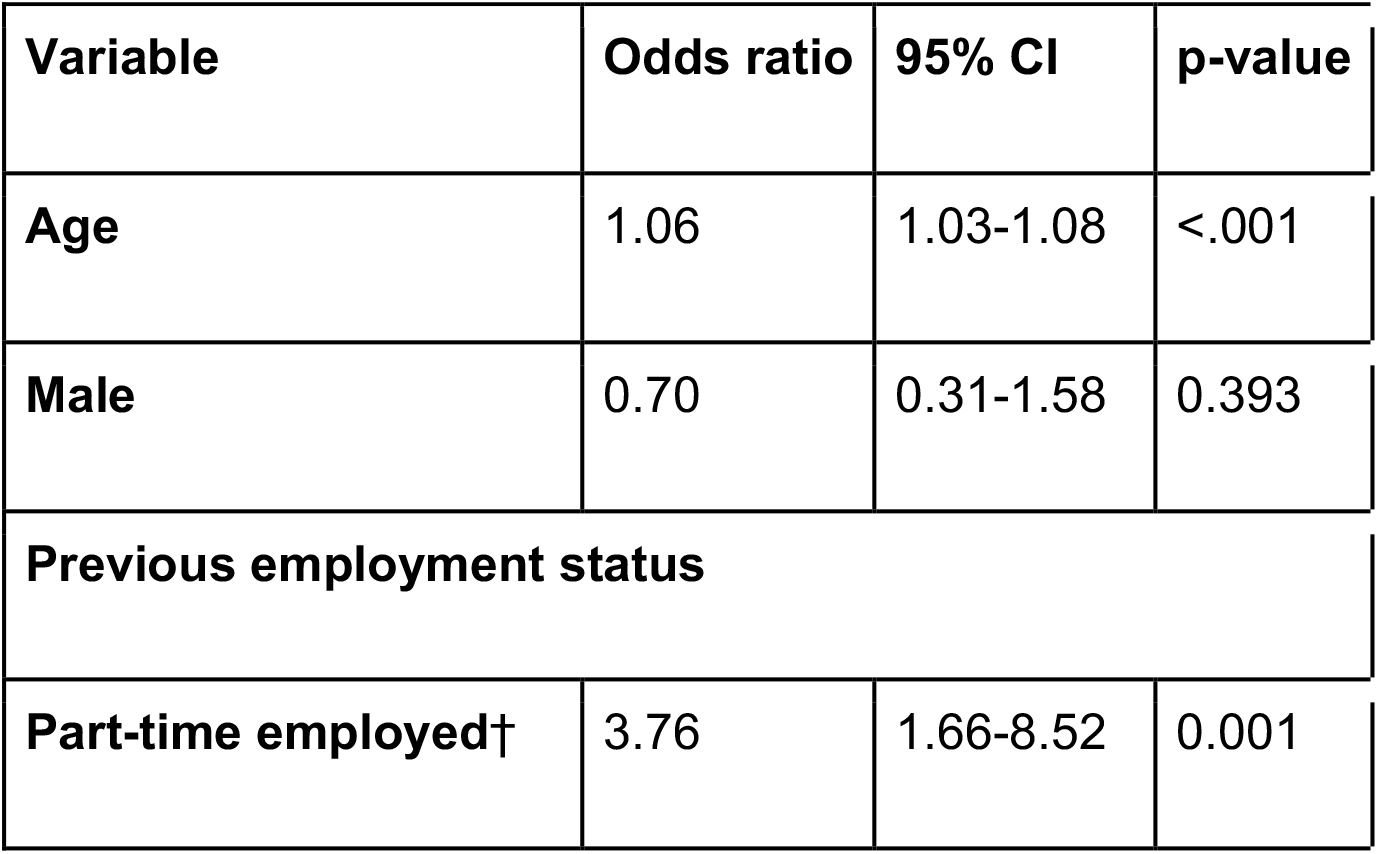
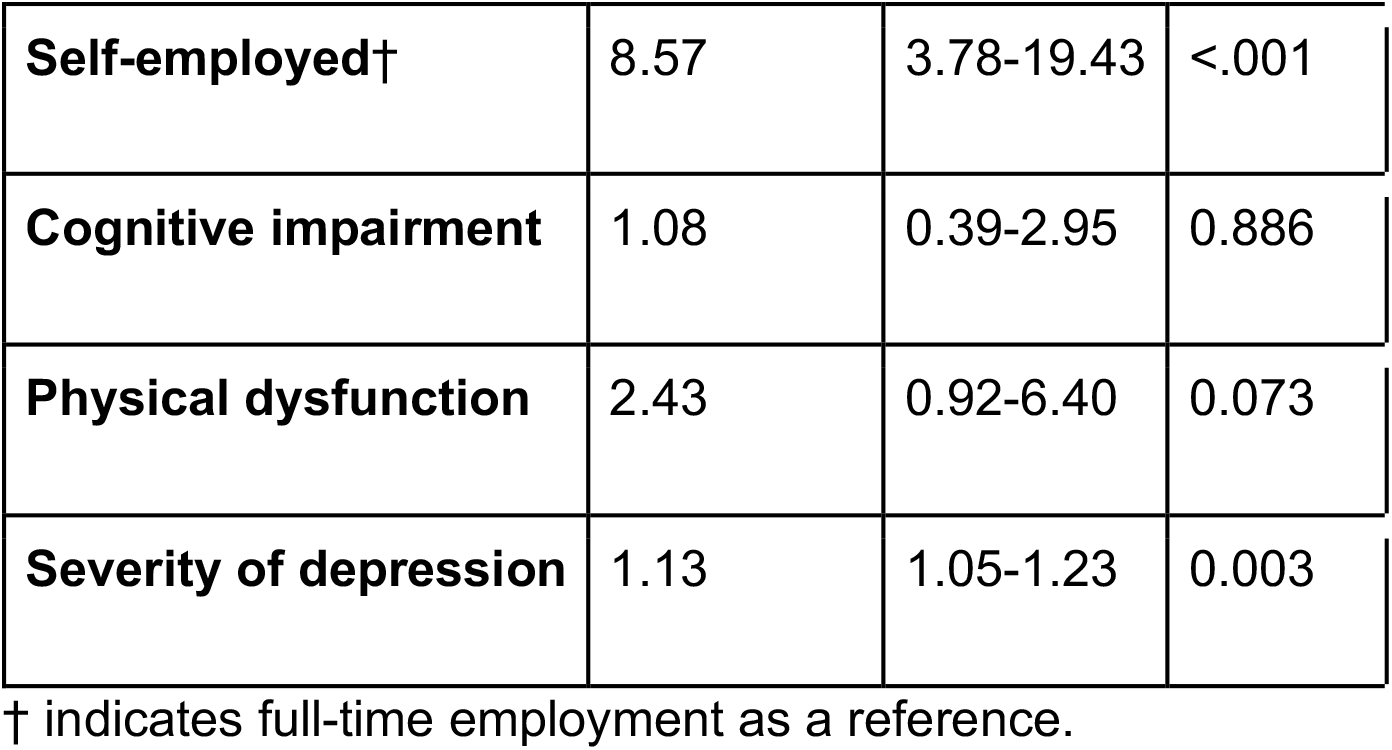
Multivariable analysis of factors associated with unemployed status 12 months after intensive care unit discharge.

### Sensitivity Analysis

First, we calculated the proportion of the unemployed after excluding all subjects who responded within the duration of the COVID-19 pandemic. The total subjects who were employed during the period prior to ICU admission was 178 and subjects who were in a state of unemployment was 40. The unemployment rate was 22.4% (95% CI, 16.9-29.2). Second, we excluded subjects who were 60 years old and above and conducted multivariable analysis as well as primary analysis. Consequently, age was not determined to be an independent factor; however, worse depressive symptoms still remained to be an independent factor among the unemployed (S2 Table).

## Discussion

We determined that one-fourth of the patients who were previously employed prior to ICU admission were subsequently unemployed at 12 months following discharge from the ICU. Additionally, over half of the unemployed patients perceived worse household financial status compared to that in the period prior to ICU admission. Severity of depressive symptoms was an independent factor for unemployment, as well as higher age and part-time or self-employed status.

On the other hand, a recent meta-analysis reported that 40% of patients did not return to work at 12 months following discharge from intensive care [1]. Second, compared with a meta-analysis [1], our findings indicate a relatively lower rate of unemployment. There are some possible reasons for the discrepancies in the findings.

First, the characteristics of patients admitted to the ICU could be contributory. Prior studies comparing ICU admissions in Japan and the U.S. found that 59.2% of patients in Japan entered the ICU after elective surgery, compared to 42.3% in the U.S., and that fewer patients entered the ICU from the emergency department in Japan [15]. Upon review, more than half of our subjects following elective surgery were included in this study.

Second, although it has not always been the case in recent years, there has been a “lifetime employment system”, which may be specific to Japan, whereas employees in their 50s, in particular, tend to be protected from redundancy [16]. This might influence our findings because a median age of 64 years old was included in our study.

Third, disability policy in each country should influence the proportion of employees returning to work among patients discharged from intensive care. A recent systematic review and meta-analysis using meta-regression revealed that disability policies in each country positively impacted the ability of ICU-discharged patients to return to work [3]. From OECD (Organization for Economic Co-operation and Development) data, the integration index that was calculated from ten criteria such as employment programs and government-provided job training was higher in Japan than the median index of OECD. [17]

Depressive symptoms are likely to be associated with unemployment status after ICU discharge. We found depressive symptoms were an independent factor associated with unemployed despite adjusting for covariates including physical function and cognitive function. These findings are consistent with our hypothesis. A previous study suggested that depression was associated with not returning to work in Australia; however, the study did not conduct multivariable analysis [18]. The present study did not indicate cause-outcome relationships between unemployment and subsequent depression. Depression may lead to unemployment and vice versa from our study design.

### Strength of the study

To date, this study is the largest multi-center study of its kind in Japan. Thus, our findings are reflective of the population admitted in Japanese ICUs.

### Limitation of the study

First, since we collected the employment status and depressive symptoms at the same time, we were not able to clarify the causal relationship between these two variables. Further research measuring depressive symptoms at the discharge from hospital or enough before collecting employment data.

Second, COVID-19 may affect our findings. For about half of the study period, Japan was under the COVID-19 pandemic; however, the level of spread was different in the region. Moreover, we conducted an ambidirectional study design; there was no effect on employment and household finance before ICU admission. Employment and household finances at 12 months following ICU discharge was likely influenced by the COVID-19 pandemic. However, we considered this influence minimal, since our sensitivity analysis showed similar results despite exclusion of subjects who responded within the COVID-19 pandemic.

Third, we included patients of all ages; thus, retirement and retirement age both affect our results. We consider this effect inconsequential to our findings based on results from our sensitivity analysis.

Fourth, we did not use validated tools to measure cognitive function. This may affect our findings; however, translated Japanese self-administered questionnaires were limited in utility.

Fifth, we excluded patients with central nervous system disease due to this study being based on self-administered questionnaires. Thus, our findings may underestimate the proportion of the unemployed.

### Implications for future research

It is likely that the proportion of those returning to work depends not only on mental health or physical function but also the employment culture and governmental policy. This study is sub-group analysis; thus, a large cohort study focusing on the return to work following intensive care admission is warranted for each country.

### Implications for clinical practice

We need to carefully monitor patients discharged from the ICU, even if they are at home. In particular, depression is associated with unemployment status and needs to be observed. The government and the local municipalities should provide medical and financial support to such patients. Additionally, community support for such patients is warranted.

## Conclusion

We found one-four patients after 12 months from intensive care were unemployed. Additionally, depressive symptoms were associated with unemployment status. employment status and mental health should be follow-up and adequate support are warranted.

## Data Availability

All data produced in the present work are contained in the manuscript

## Data availability statement

pAll relevant data are within the manuscript and Supporting Information files.

## Acknowledgements

The following SMAP-HoPe Study Project investigators were involved in the protocol: Ryuta Indo, Hiroomi Tatsumi, Atsuko Handa, Kazuyo Koori, Ayano Kudo, Kayo Kitaura, Etsuko Moro, Shin Nunomiya, Akira Ouchi, Masako Sato, Yoshiaki Inoue, Etsuko Tsukioka, Yasuhiro Kishi, Chiaki Fujii, Kohei Matsuba, Hiroki Isonishi, Ikumi Kobashi, Miki Toyoshima, Masahiro Yamane, Yumi Kajiyama, and Yoshifumi Heshiki

**S1 Table 1. Distribution of age group and proportion of unemployed subjects stratified by age group**

**S2 Table 2. Sensitivity analysis**: Multivariable analysis of factors associated with unemployed status 12 months after intensive care unit discharge among patients after excluding patients aged 60 years or older

## References

1. Kamdar BB, Suri R, Suchyta MR, Digrande KF, Sherwood KD, Colantuoni E, et al. Return to work after critical illness: a systematic review and meta-analysis. Thorax. 2020;75: 17–27. doi:10.1136/thoraxjnl-2019-213803

2. Griffiths J, Hatch RA, Bishop J, Morgan K, Jenkinson C, Cuthbertson BH, et al. An exploration of social and economic outcome and associated health-related quality of life after critical illness in general intensive care unit survivors: a 12-month follow-up study. Crit Care. 2013;17: R100. doi:10.1186/cc12745

3. Su H, Dreesmann NJ, Hough CL, Bridges E, Thompson HJ. Factors associated with employment outcome after critical illness: Systematic review, meta-analysis, and meta-regression. J Adv Nurs. 2021;77: 653–663. doi:10.1111/jan.14631

4. Kawakami D, Fujitani S, Morimoto T, Dote H, Takita M, Takaba A, et al. Prevalence of post-intensive care syndrome among Japanese intensive care unit patients: a prospective, multicenter, observational J-PICS study. Crit Care. 2021;25: 69. doi:10.1186/s13054-021-03501-z

5. Myhren H, Ekeberg Ø, Stokland O. Health-related quality of life and return to work after critical illness in general intensive care unit patients: a 1-year follow-up study. Crit Care Med. 2010;38: 1554–1561. doi:10.1097/CCM.0b013e3181e2c8b1

6. Rothenhäusler H-B, Ehrentraut S, Stoll C, Schelling G, Kapfhammer H-P. The relationship between cognitive performance and employment and health status in long-term survivors of the acute respiratory distress syndrome: results of an exploratory study. Gen Hosp Psychiatry. 2001;23: 90–96. doi:10.1016/s0163-8343(01)00123-2

7. Zisopoulos G, Roussi P, Mouloudi E. Psychological morbidity a year after treatment in intensive care unit. Health Psychol Res. 2020;8: 8852. doi:10.4081/hpr.2020.8852

8. Norman BC, Jackson JC, Graves JA, Girard TD, Pandharipande PP, Brummel NE, et al. Employment Outcomes After Critical Illness. Crit Care Med. 2016;44: 2003–2009. doi:10.1097/ccm.0000000000001849

9. Vlasveld MC, van der Feltz-Cornelis CM, Bültmann U, Beekman ATF, van Mechelen W, Hoedeman R, et al. Predicting return to work in workers with all-cause sickness absence greater than 4 weeks: a prospective cohort study. J Occup Rehabil. 2012;22: 118–126. doi:10.1007/s10926-011-9326-0

10. Unoki T, Sakuramoto H, Uemura S, Tsujimoto T, Yamaguchi T, Shiba Y, et al. Prevalence of and risk factors for post-intensive care syndrome: Multicenter study of patients living at home after treatment in 12 Japanese intensive care units, SMAP-HoPe study. PLoS One. 2021;16: e0252167. doi:10.1371/journal.pone.0252167

11. Herdman M, Gudex C, Lloyd A, Janssen M, Kind P, Parkin D, et al. Development and preliminary testing of the new five-level version of EQ-5D (EQ-5D-5L). Qual Life Res. 2011;20: 1727–1736. doi:10.1007/s11136-011-9903-x

12. Zigmond AS, Snaith RP. The Hospital Anxiety and Depression Scale. Acta Psychiatr Scand. 1983;67: 361–370. doi:10.1111/j.1600-0447.1983.tb09716.x PMID -6880820

13. Hatta H, Higashi A, Yashiro H, Kotaro O, Hayashi K, Kiyota K, et al. A Validation of the Hospital Anxiety and Depression Scale. Jpn J Psychosom Med. 1998;38: 309–315. doi:10.15064/jjpm.38.5_309

14. Okamoto S, Okamura T, Komamura K. Employment and health after retirement in Japanese men. Bull World Health Organ. 2018;96: 826–833. doi:10.2471/BLT.18.215764

15. Sirio CA, Tajimi K, Taenaka N, Ujike Y, Okamoto K, Katsuya H. A Cross-Cultural Comparison of Critical Care Delivery Japan and the United States. Chest. 2002;121: 539–548. doi:10.1378/chest.121.2.539

16. Kambayashi R, Kato T. Long-Term Employment and Job Security over the Past 25 Years: A Comparative Study of Japan and the United States. ILR Review. 2017;70: 359–394. doi:10.1177/0019793916653956

17. Organisation for Economic Co-operation and Development. Sickness, disability and work breaking the barriers : a synthesis of findings across OECD countries. Paris: OECD; 2010. Available: https://www.worldcat.org/title/sickness-disability-and-work-breaking-the-barriers-a-synthesis-of-findings-across-oecd-countries/oclc/1136220722?referer=di&ht=edition

18. Hodgson CL, Haines KJ, Bailey M, Barrett J, Bellomo R, Bucknall T, et al. Predictors of return to work in survivors of critical illness. J Crit Care. 2018;48: 21–25. doi:10.1016/j.jcrc.2018.08.005

