## Supplementary Table 1 for "Employment status and its associated factor for patients 12 months after intensive care: Secondary analysis of the SMAP-HoPe-study"

**S1 Table**

**Distribution of age group and proportion of unemployed subjects stratified by age group**

| **Age** | **n (%)** | **Proportion of unemployed, n (%)** |
| --- | --- | --- |
| **<19** | 1 (0.3) | 0 |
| **20-29** | 10 (3.1) | 2 (20.0) |
| **30-39** | 16 (4.9) | 2 (12.5) |
| **40-49** | 39 (11.9) | 4 (10.3) |
| **50-59** | 59 (18.0) | 8 (13.6) |
| **60-69** | 93 (28.4) | 25 (26.9) |
| **70-79** | 91 (27.7) | 33 (36.3) |
| **>80** | 19 (5.8) | 5 (26.3) |
| **Total** | 328 (100) | 79 (24.1) |
