## Supplementary Table 2 for "Employment status and its associated factor for patients 12 months after intensive care: Secondary analysis of the SMAP-HoPe-study"

**S2 Table. Sensitivity analysis**

Multivariable analysis of factors associated with unemployed status 12 months after intensive care unit discharge among patients after excluding patients aged 60 years or older

| **Variable** | **Odds Ratio** | **95% CI** | **p** |
| --- | --- | --- | --- |
| **Age** | 1.04 | 0.99-1.10 | 0.129 |
| **Male** | 0.93 | 0.31-2.76 | 0.898 |
| **Previous employment status** | | | |
| **Part-time employed** | 5.97 | 2.41-14.77 | <.001 |
| **Self-employed** | 7.73 | 3.15-18.96 | <.001 |
| **Cognitive impairment** | 0.79 | 0.20-3.08 | 0.731 |
| **Physical dysfunction** | 1.90 | 0.57-6.26 | 0.294 |
| **Severity of depression** | 1.17 | 1.06-1.30 | 0.002 |

CI, Confidence Interval
